# Emerging viruses are an underestimated cause of undiagnosed febrile illness in Uganda

**DOI:** 10.1101/2023.04.27.23288465

**Authors:** Shirin Ashraf, Hanna Jerome, Daniel Lule Bugembe, Deogratius Ssemwanga, Timothy Byaruhanga, John Timothy Kayiwa, Robert Downing, Jesus F. Salazar-Gonzalez, Maria G. Salazar, James G. Shepherd, Chris Davis, Nicola Logan, Sreenu B. Vattipally, Gavin S. Wilkie, Ana da Silva Filipe, Alfred Ssekagiri, Prossy Namuwulya, Henry Bukenya, Brian K. Kigozi, Weronika Witkowska McConnell, Brian J. Willett, Stephen Balinandi, Julius Lutwama, Pontiano Kaleebu, Josephine Bwogi, Emma C. Thomson

**Author notes:** Contributed equally (Shirin Ashraf, Hanna Jerome). Contributed equally (Josephine Bwogi, Emma C. Thomson).

## Abstract

**Background:** Viruses that cause acute febrile illness (AFI) in sub-Saharan Africa cause a spectrum of disease from mild to life-threatening. Viral infection is often undiagnosed, as routine diagnostics are insufficient to capture the diversity of circulating pathogens.

**Methods:** 1281 patients with fever of 2-7 days were prospectively recruited from three sites in Uganda as part of the CDC-UVRI AFI Study and screened with enhanced diagnostics. Plasma from 233 undiagnosed patients was analysed using metagenomic next-generation sequencing (mNGS). Confirmatory testing was carried out by PCR and serology.

**Findings:** Thirty-eight viral pathogens were identified by mNGS in 35/233 (15%) undiagnosed patients including Measles, Hepatitis A/B/E viruses, Human immunodeficiency virus-1, Rhinovirus, Rotavirus-like virus, Human herpesvirus 6B, Human parainfluenza virus 3 and Enteroviruses. Four high-consequence arboviruses were found in six patients; Crimean-Congo haemorrhagic fever virus, Rift Valley fever virus, dengue virus and yellow fever virus. Le Dantec virus, last reported in 1969, was detected and confirmed by serology in one patient (and a contact of that patient). The majority of patients (23/30; 76%) diagnosed with an acute viral infection were treated with antibiotics and/or (12/30; 40%) antimalarials.

**Interpretation:** AFI in Uganda is commonly associated with undiagnosed viral infection, including high-consequence and rarely reported viruses. This highlights an ongoing risk to public health and the need for improved vigilance. MNGS alongside diagnostic serology is a powerful method for clinical surveillance to investigate circulating viral pathogens. Cost-effective diagnostic assays should be adapted according to regional needs for testing.

**Funding:** Medical Research Council and Wellcome Trust

## INTRODUCTION

East Africa is a biodiverse region and a hotspot for viral zoonoses. In the last decade, Uganda has reported outbreaks of haemorrhagic fever caused by Ebola (EBOV), Sudan (SUDV), Marburg (MARV), Rift Valley fever (RVFV), yellow fever (YFV) and Crimean-Congo haemorrhagic fever (CCHFV) viruses^1^. Many vector-borne viral infections are prevalent in Uganda, and several were described here for the first time, including Zika virus (ZIKV), Semliki Forest virus (SFV), chikungunya virus (CHIKV), O’Nyong Nyong virus (ONNV) and West Nile virus (WNV)^2^. Anthropogenic change, as a result of frequent travel, increasing commerce, and shifting global climate patterns raises the risk of spread within and outside zoonotic hotspots^3^. Many infections are likely missed due to the scarcity of diagnostic tests, increasing the risk of spread of high consequence viruses. Further, they may be misattributed to malaria or bacterial infection, increasing the risk of antimicrobial resistance. In this study, we investigated the occurrence of undiagnosed viral infections as a cause of acute febrile illness (AFI).

AFI is a common cause of presentation to healthcare centres (HC) across sub-Saharan Africa (SSA), despite a significant reduction in malaria prevalence following successful intervention campaigns^4^. Communicable diseases are estimated to account for up to 36% productivity losses in SSA^5^. The contribution of viral infection to AFI in Africa is at least 8-60%, likely an under-estimate as population studies have relied on traditional diagnostic tests which are limited to selected pathogens^6–9^. A thorough understanding of the burden of viral disease is essential for implementing prevention strategies by limiting vector-borne transmission, introducing vaccination where needed and available, rapid cost-effective diagnostics and treatments. Metagenomic next-generation sequencing (mNGS) allows for unbiased identification of viral genomes in clinical samples, including known and novel pathogens. In this study, we used mNGS to identify viral infections in patients in Uganda presenting with AFI that remained undiagnosed after routine serological and PCR-based assays.

## METHODS

### Patients and sampling

1281 patients were recruited prospectively with informed consent into the AFI study from three study sites in Uganda and assigned an anonymized study number, not known to anyone outside the recruitment team at each healthcare provision site; Ndejje HC IV (Wakiso), St. Paul’s HC IV (Kasese), and Adumi HC IV (Arua) between April 2011 and January 2013 (**Figure 1a**). Inclusion criteria were >2 years age with (a) a reported fever lasting 2-7 days or ≥38°C temperature on admission, or (b) symptoms consistent with brucellosis or typhoid fever. Cases with clinical evidence of an alternative diagnoses such as otitis media were excluded. Samples were obtained at presentation (acute sample) and 14-21 days later (convalescent sample). Diagnostic assays including serology, blood culture and blood films were carried out to identify and exclude active or recent infection with malaria, typhoid, leptospirosis, rickettsiae, CHIKV, dengue (DENV), WNV, YFV and ONNV. 210 patients remained undiagnosed and acute plasma were retrospectively tested using mNGS. In addition, 23 samples were obtained from two outbreaks of febrile illness that occurred within the time period of the study.

**Figure 1.**
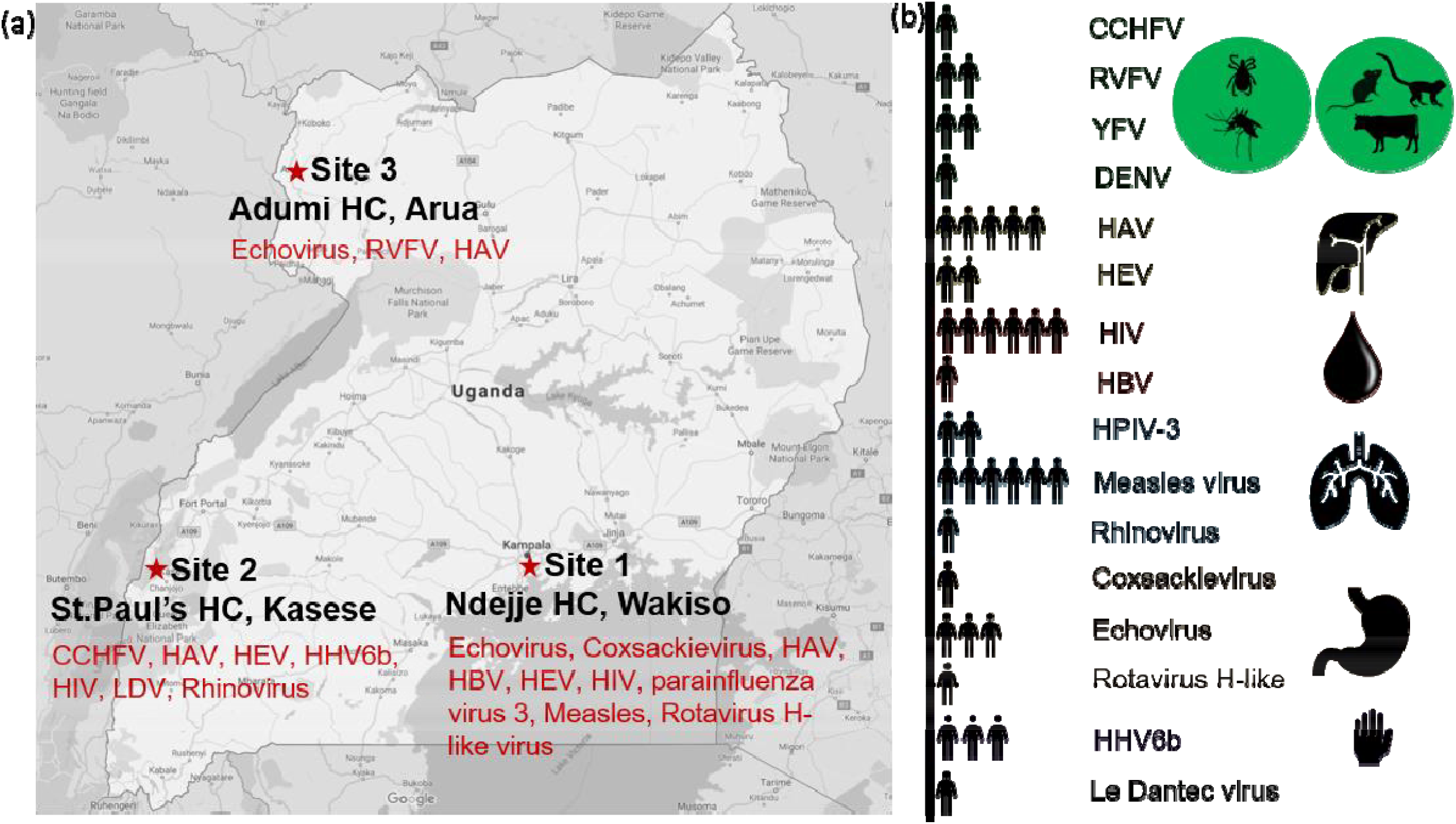
Undiagnosed viruses identified in the AFI study. (a) AFI study sites and locations. Viruses identified from undiagnosed samples are listed by site. (b) Viruses detected by mNGS grouped as zoonoses/arboviruses (green), hepatitis (yellow), blood-borne infections (red), respiratory viruses (blue), gastrointestinal infections (brown), viruses associated with rash (purple) and uncharacterized (grey). Icons show the number of patients infected with each virus.

### Ethics statement

Ethical approval for the AFI study was granted by the Uganda Virus Research Institute (UVRI) Research Ethics Committee (GC/127/10/02/19) and the Uganda National Council for Science and Technology (HS767).

### Sample processing and mNGS

RNA was extracted from 200μl of plasma using the Agencourt RNAdvance Blood Kit (Beckman Coulter), according to manufacturer’s instructions, including DNase treatment at 37°C for 15min. RNA was reverse transcribed using Superscript III (Invitrogen) followed by dsDNA synthesis with NEBNext Enzyme (New England Biolabs). DNA libraries were prepared using LTP low-input Library preparation kit (KAPA Biosystems) as previously described^10^. Resulting libraries were quantified with the Qubit 3.0 fluorometer (Invitrogen) and their size determined using a 4200 TapeStation (Agilent). Libraries were pooled in an equimolar ratio and sequenced on the Illumina MiSeq and NextSeq platforms.

### Bioinformatic analysis

Raw Fastq files were searched directly for evidence of viral sequences using Diamond BLASTX (**Figure S1a**)^11^. *De novo* assembly was carried out using dipspades and contigs were identified using Diamond BLASTX against the nr database^12^. Viral hits detected with blastx were confirmed with blastn and mapped to the closest reference genome using *Tanoti*, a mapper developed for use with highly diverse viral genomes (github.com/vbsreenu/Tanoti). A diagnosis threshold of 28 nucleotides was estimated as a reliable cut-off following an analysis of all common human viral pathogens and removal of genomic repeat regions, well under the minimum sequence length in our samples (**Figure S1b**). Maximum-likelihood phylogenetic analysis was carried out using IQTREE and 1000 ultra-fast bootstrap replicates using relevant reference sequences^13^. Uncorrected pairwise-distances were estimated using MEGA 10.0^14^.

### Capture ELISA

Rhabdovirus (Le Dantec; LDV and Adumi virus) glycoprotein genes (synthesized by Eurofins Genomics) excluding the transmembrane regions (identified using TMHMM and TMpred) were amplified and cloned into the secretory mammalian expression vector pHLSec containing a C-terminal 6xHistidine tag^15,16^. Human embryonic kidney cells (HEK 293T) were maintained in Dulbecco’s modified Eagle’s medium (DMEM) supplemented with 100IU/ml penicillin, 100μg/ml streptomycin, 2mM glutamine and 10% foetal bovine serum. Cells were transfected (Fugene 6, Promega) and cell supernatant containing secreted recombinant glycoproteins was harvested at 48-hours and expression confirmed by Western Blotting against 6xHis. ELISA plates (ImmulonHB) were coated with rabbit anti-His antibody overnight and blocked with 2.5% BSA/PBS at 37°C for 1 hour. Cell supernatant containing glycoprotein was added to the wells followed by patient serum, both for 1 hour at 37°C. This was followed by HRP-conjugated goat anti-human IgG at room temperature for 1 hour. Wells were washed after each step with 0.1% Tween-20/PBS. Reactions were developed using TMB substrate, stopped with 0.16M sulphuric acid and read on a spectrophotometer (Pherastar).

### Pseudotype neutralization assay

The use of Vesicular Stomatitis Virus (VSV) in which the glycoprotein sequence has been replaced (VSVΔG*luc*) to generate pseudotyped viruses has been described previously^17^. Glycoprotein genes were cloned into the eukaryotic expression vector and transfected into HEK-293T cells using polyethylenimine followed by infection with VSVΔG*luc*-VSV-G at a multiplicity of infection of 0.02. Cells were incubated for one hour at 37°C, washed with PBS and re-incubated in DMEM. Supernatants were harvested 72-hours post infection. Pseudoparticle activity was assessed on HEK-293T cells by measuring luciferase activity at 72-hours on a Microbeta 1450 Jet luminometer (Perkin Elmer).

For neutralisation assays, 1×10^4^ HEK-293T cells were plated in a white 96-well plate and incubated at 37°C for one hour. Four-fold serum dilutions were prepared in DMEM ranging from 1:8 to 1:131072. Serum was mixed with either VSVΔG*luc*-VSV-G or VSVΔG*luc*-LDV-G at a concentration expected to generate a luciferase reading of 1 × 10^6^ counts per second and incubated for 1 hour at 37°C. 50μl per well of the pseudotype/serum mixture was added to the cells and incubated for 24 hours. Luciferase substrate was added to each well (Steadylite plus™, Perkin Elmer) and luminescence measured on a Chameleon V plate scintillation counter (Hidex Oy).

## RESULTS

### Demographics and clinical analysis

15% of patients with undiagnosed AFI (35/233) had evidence of acute viral infection by mNGS (**Figure 1b**). Demographic and clinical data was available for 30/35 patients with AFI. The median age at clinical presentation was 7.5 years (range 2-40). 80% of cases occurred in children under the age of 16. Males were more commonly represented than females (20/30; 67% versus 10/30; 33%). The majority had been recently bitten by mosquitoes (22/30; 73%) and/or other arthropod vectors, including biting flies (3/30; 10%), ticks (3/30; 10%), fleas (2/30; 7%) and lice (1/30; 3%). Ten percent reported haemorrhagic symptoms but none of these had evidence of a viral haemorrhagic fever (VHF) virus by mNGS. At presentation, the median temperature was 38.2°C. Other clinical symptoms are described in **Table 1**. Common presumptive diagnoses included malaria, respiratory tract infection, typhoid fever and unknown. The majority 23/30 (77%) of patients were presumptively treated with antibiotics, 12 (40%) were treated with antimalarials and 5 (17%) with antihelminth therapy.

**Table 1.**
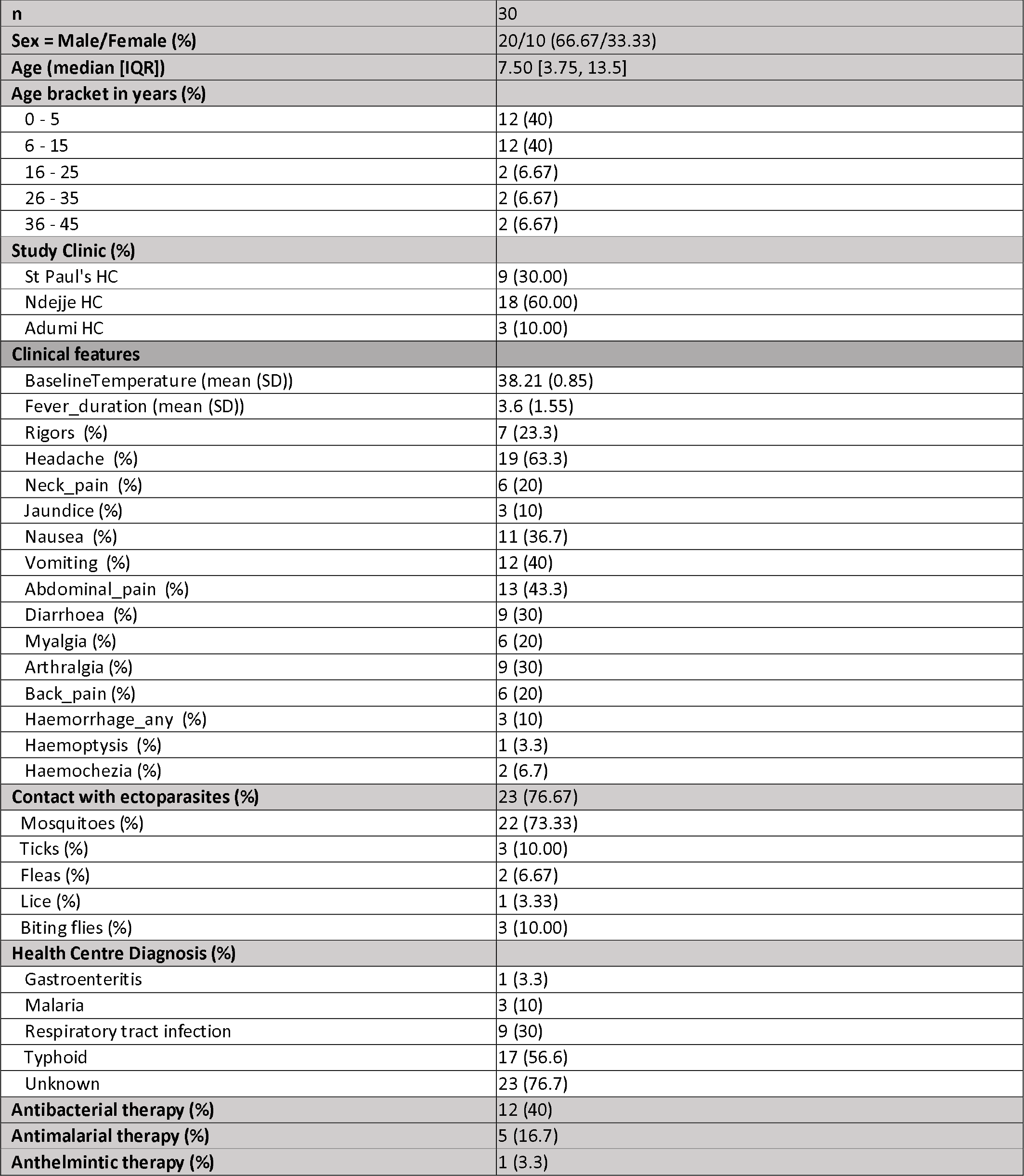
Demographics and clinical characteristics of patients diagnosed with viral infections.

Of the 35 patients diagnosed by mNGS, 6 (17%) had a virus associated with VHF (CCHFV, RVFV, YFV and DENV), 9 (25%) with viruses associated with respiratory infection (human parainfluenza 3, rhinovirus C and measles virus), 8 (22%) with hepatitis causing viruses (HAV, HBV, HEV), and 5 (14%) with gastroenteritis associated viruses (**Table 2, Supplementary Table S1**). Six patients (17%) were infected with human immunodeficiency virus-1 (HIV-1). One patient was infected with the rhabdovirus LDV. Two of these cases, one each of yellow fever and RVF were correctly identified in the context of a known outbreak.

**Table 2.**
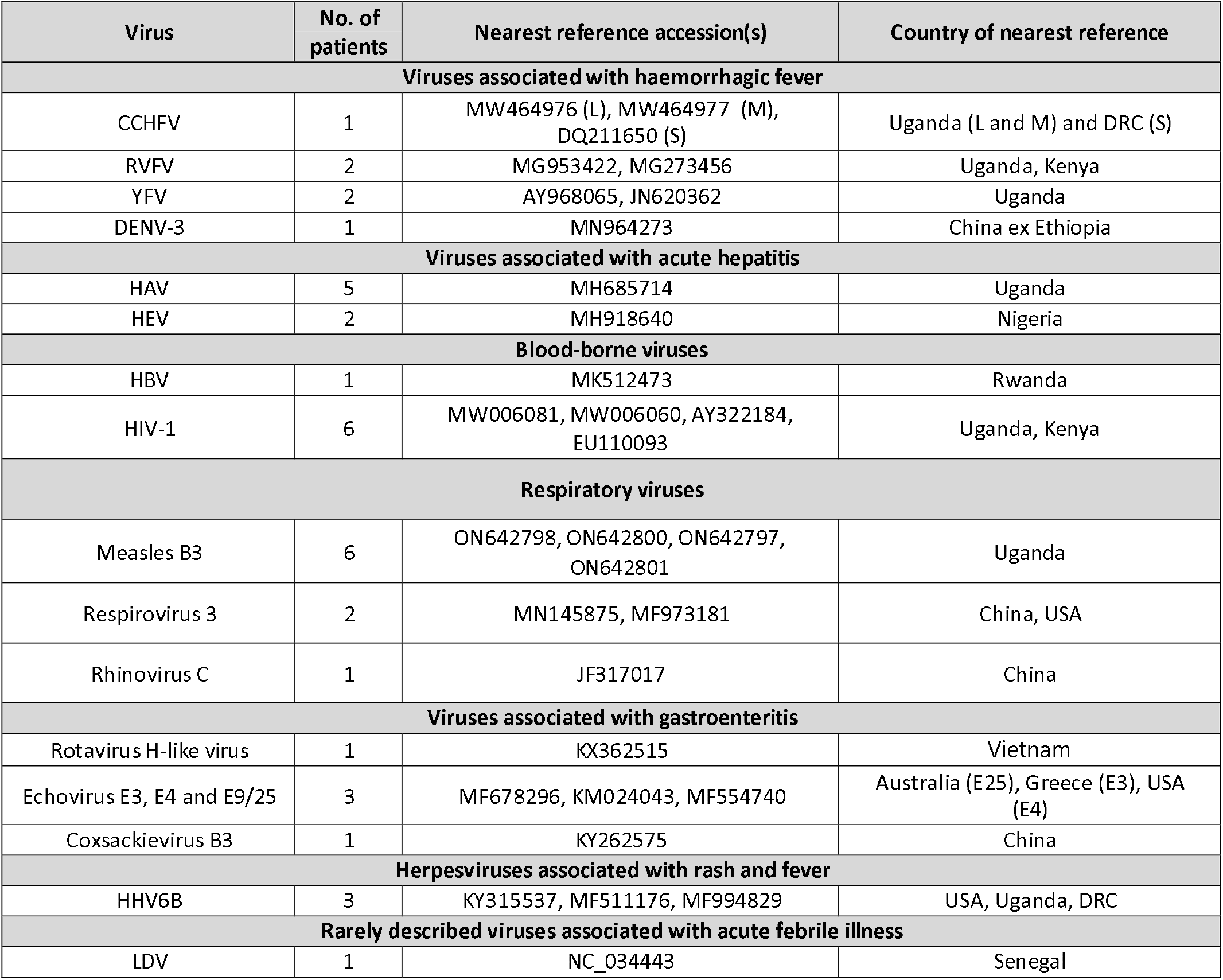
Viruses detected by mNGS in undiagnosed patients in the AFI study.

| Virus | No. of patients | Nearest reference accession(s) | Country of nearest reference |
| --- | --- | --- | --- |
| **Viruses associated with haemorrhagic fever** |  |  |  |
| CCHFV | 1 | MW464976 (L), MW464977 (M), DQ211650 (S) | Uganda (L and M) and DRC (S) |
| RVFV | 2 | MG953422, MG273456 | Uganda, Kenya |
| YFV | 2 | AY968065, JN620362 | Uganda |
| DENV-3 | 1 | MN964273 | China ex Ethiopia |
| **Viruses associated with acute hepatitis** |  |  |  |
| HAV | 5 | MH685714 | Uganda |
| HEV | 2 | MH918640 | Nigeria |
| **Blood-borne viruses** |  |  |  |
| HBV | 1 | MK512473 | Rwanda |
| HIV-1 | 6 | MW006081, MW006060, AY322184, EU110093 | Uganda, Kenya |
| **Respiratory viruses** |  |  |  |
| Measles B3 | 6 | ON642798, ON642800, ON642797, ON642801 | Uganda |
| Respirovirus 3 | 2 | MN145875, MF973181 | China, USA |
| Rhinovirus C | 1 | JF317017 | China |
| **Viruses associated with gastroenteritis** |  |  |  |
| Rotavirus H-like virus | 1 | KX362515 | Vietnam |
| Echovirus E3, E4 and E9/25 | 3 | MF678296, KM024043, MF554740 | Australia (E25), Greece (E3), USA (E4) |
| Coxsackievirus B3 | 1 | KY262575 | China |
| **Herpesviruses associated with rash and fever** |  |  |  |
| HHV6B | 3 | KY315537, MF511176, MF994829 | USA, Uganda, DRC |
| **Rarely described viruses associated with acute febrile illness** |  |  |  |
| LDV | 1 | NC_034443 | Senegal |

### Viruses associated with VHF

Genomes from four VHF-associated viruses were detected in six febrile patients; CCHFV (patient 138-2), RVFV (patients 117-3, RVF58), DENV (patient ARB248) and YFV (patients YFV685, ARB176).

The CCHF patient (138-2) was a male in his 30s from Kasese district. He presented with fever (37.9°C), headache, nausea and joint pain. He had been bitten by mosquitoes but had not been aware of tick bites and had not recently slaughtered animals. He was treated with vitamins and paracetamol and recovered. Partial (10-25% coverage) CCHFV sequences across all three segments; large (L), medium (M) and small (S) were detected by mNGS (**Figure 2a**). The L and S segments had >90% nucleotide identity to previously identified genomes from Uganda and Democratic Republic of the Congo, respectively (**Table 2, S1**). The M segment had 81% nucleotide identity to a genome from Uganda, suggestive of higher divergence or possible reassortment (**Figure 2d**). Convalescent serology was not available from the patient, who recovered and had moved abroad.

**Figure 2.**
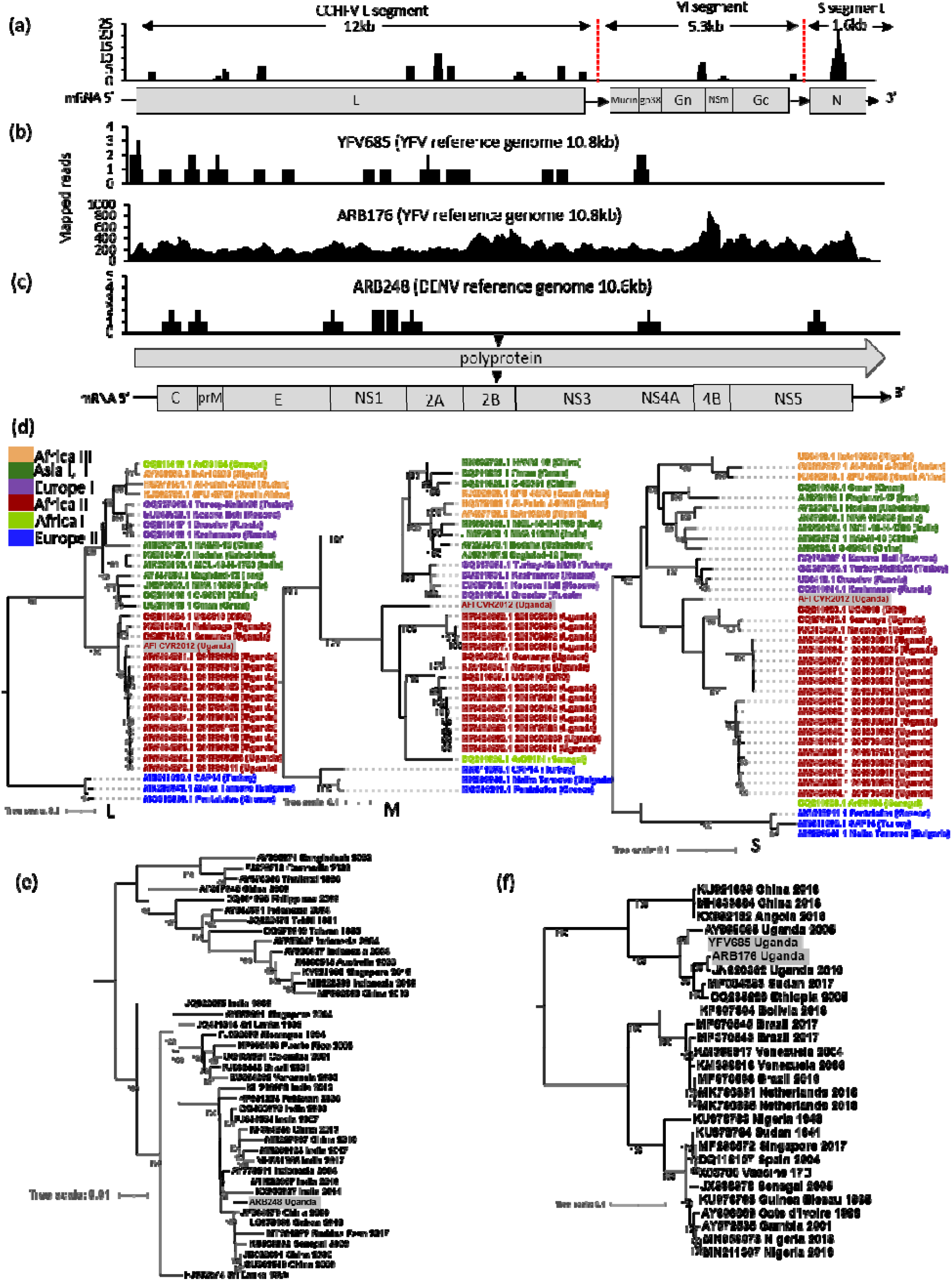
Viruses associated with haemorrhagic fever. (a-c) Coverage plots for CCHFV, YFV, DENV identified by mNGS showing number of reads (y-axis) at each nucleotide position across the genome (x-axis) for each virus. Reference orthonairovirus and flavivirus genomes indicate regions covered by sequencing reads. (d) Phylogenetic tree of full-length nucleotide sequence of CCHFV showing placement of patient isolate for L, M, S segments. Tip labels show strain name and country of isolation. Colours indicate geographically distributed lineages of CCHFV. (e) Full genome nucleotide tree of DENV genotype 3 sequences showing country and year of isolation. (f) Full genome nucleotide tree of YFV sequences showing country and year of isolation. Evolutionary scale is showing below for each tree. Ultrafast bootstrap values >90 are shown for nodes. Patient samples are highlighted in grey. Substitution models used: GTR+F+I+G4 (CCHF-L, DENV-3), GTR+F+R3 (CCHF-M), TIM2+F+I+G4 (CCHF-S), GTR+F+R2 (YFV).

Two cases of RVF (117-3, RVF58) were detected. RVF58 was suspected due to occupational risk (contact with slaughtered animals) and suggestive symptoms during an outbreak, and was confirmed by serology. 117-3 was a woman in her 30s from a rural part of Arua district who gave a 3-day history of fever but whose temperature was recorded as 36.3°C on admission. She described chills, headache, eye, neck and joint pain, nausea, diarrhoea and jaundice. She had been bitten by mosquitoes and fleas. She was given paracetamol and attended a traditional healer for further treatment. Convalescent serology was not available for this patient. Viral reads most closely matched the L segment of a Ugandan strain in case 1 (RVF58) and the M segment of a Kenyan strain in case 2 (117-3).

Two cases of YFV infection and one DENV infection (genotype 3) were detected during suspected YFV outbreaks and confirmed by convalescent serology. All genomes identified by mNGS were related to East/Central Africa lineages (**Table 2, S1; Figure 2 b,c,e,f**).

While haemorrhagic symptoms were recorded for 3/30 (10%) AFI study cohort patients, none of the these had a known VHF virus detected. The study patients described above with CCHFV and RVFV did not report classical haemorrhagic symptoms.

### Le Dantec virus

Patient 220-2 was a male aged between 5 and 10 years from Kasese district. He presented to hospital with a 38.5°C fever of 4 days, with headache, abdominal pain, vomiting and joint pains. He reported having been bitten by mosquitoes. He was treated with ciprofloxacin and paracetamol for suspected typhoid fever and made a full recovery. By mNGS, we detected the full genomic sequence of a previously rarely reported rhabdovirus; Le Dantec virus (LDV; **Figure 3a**). This was confirmed by direct RT-PCR from patient plasma and Sanger sequencing. The LDV genome had 94% nucleotide identity to the RefSeq LDV genome (NC_034443.1), isolated for the first time in 1965 in Senegal and sequenced from an archival sample in 2015^18^ (**Figure 3d**). In keeping with an active infection, we demonstrated immunological response to LDV glycoprotein by ELISA and pseudotype neutralisation assay, in both the patient and in a contact of the patient who had been unwell at the same time, using convalescent serum (**Figure 3b,c**).

**Figure 3.**
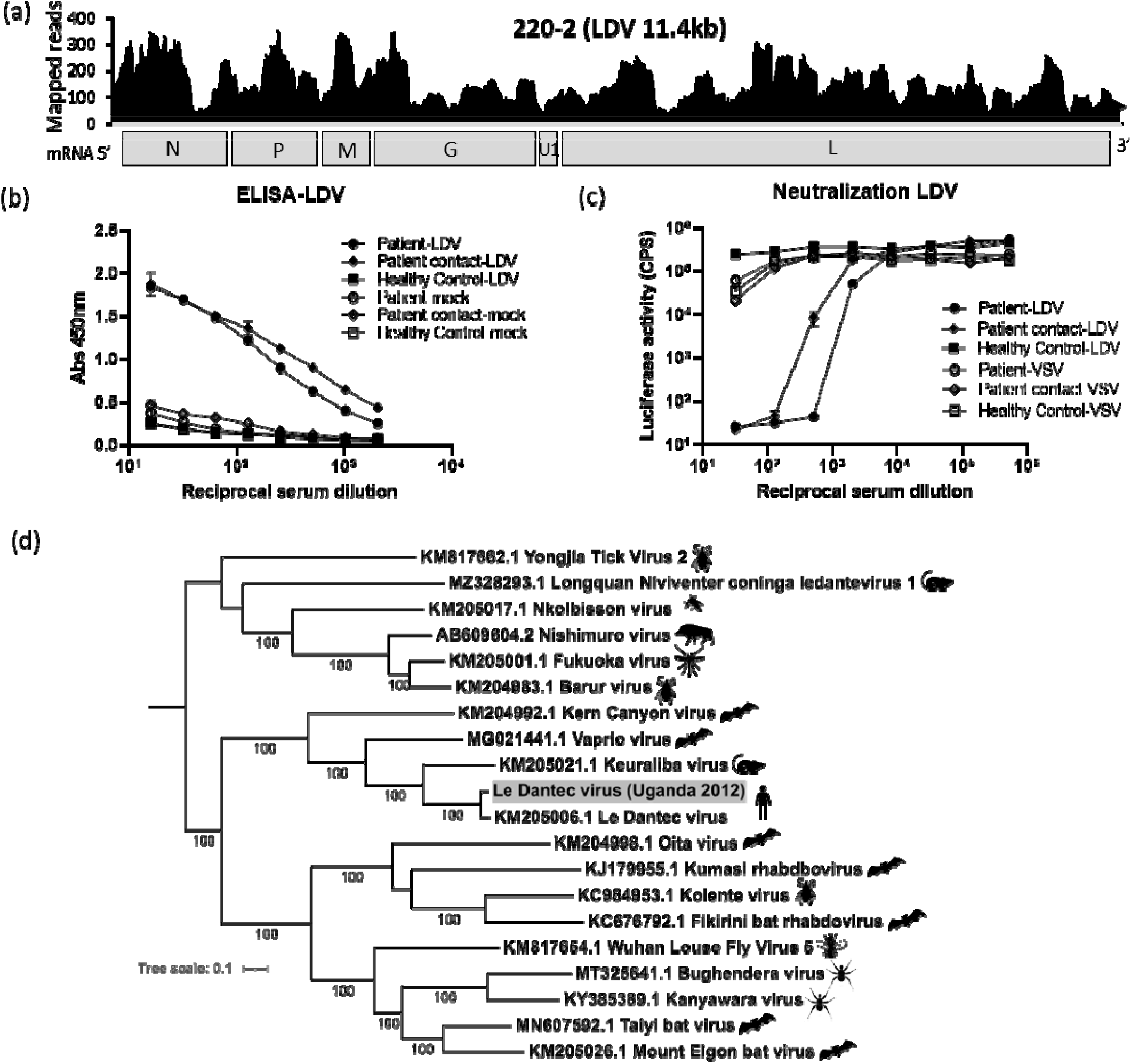
Le Dantec virus (LDV). (a) Coverage plots for LDV identified by mNGS showing number of reads (y-axis) at each nucleotide position across the genome (x-axis) for each virus. Sequencing depth for different genes across the genome is shown. (b) ELISA analysis showing IgG antibody response to recombinant LDV glycoprotein in patient 220-2, contact of patient and a healthy individual from Uganda. Plasmid containing an Ephrin B2 insert was used as a mock recombinant protein. The plot shows absorbance OD values at 450nm (y-axis) against increasing serum dilutions (x-axis). (c) Pseudotype neutralisation assay showing luciferase reporter activity (y-axis) against increasing serum dilutions (x-axis) for patient 220-2, contact and healthy serum using LeDantec pseudotype virus compared with VSV control. (d) Full genome maximum likelihood phylogenetic nucleotide tree using the GTR+F+R4 model for ledantevirus sequences illustrating the reported host for each virus. Evolutionary scale and ultrafast bootstrap values >90 are shown. The patient sample is highlighted in grey.

### Viruses associated with acute hepatitis

Hepatitis A (HAV) and hepatitis E (HEV) viruses were detected in 5 (patients ARB169, 218-1, 169-1, 184-2, 145-3) and 2 (patients 20-2, 60-1) patients respectively. Clinical data was available for four of the HAV cases. The age of infection ranged from 2 to 15 years. All were febrile at presentation. Three of four patients presented with vomiting, one had diarrhoea and one had jaundice. All HAV genomes were genotype IB and related to previously reported sequences from Uganda (**Figure 4a, S3e**). The HEV patients were aged between 0 and 5 years, and febrile at presentation. One had diarrhoea and vomiting and the other had undifferentiated fever. The two HEV isolates were genotype 1e and closely related to a Nigerian sequence (**Figure 4b, S3d**).

**Figure 4.**
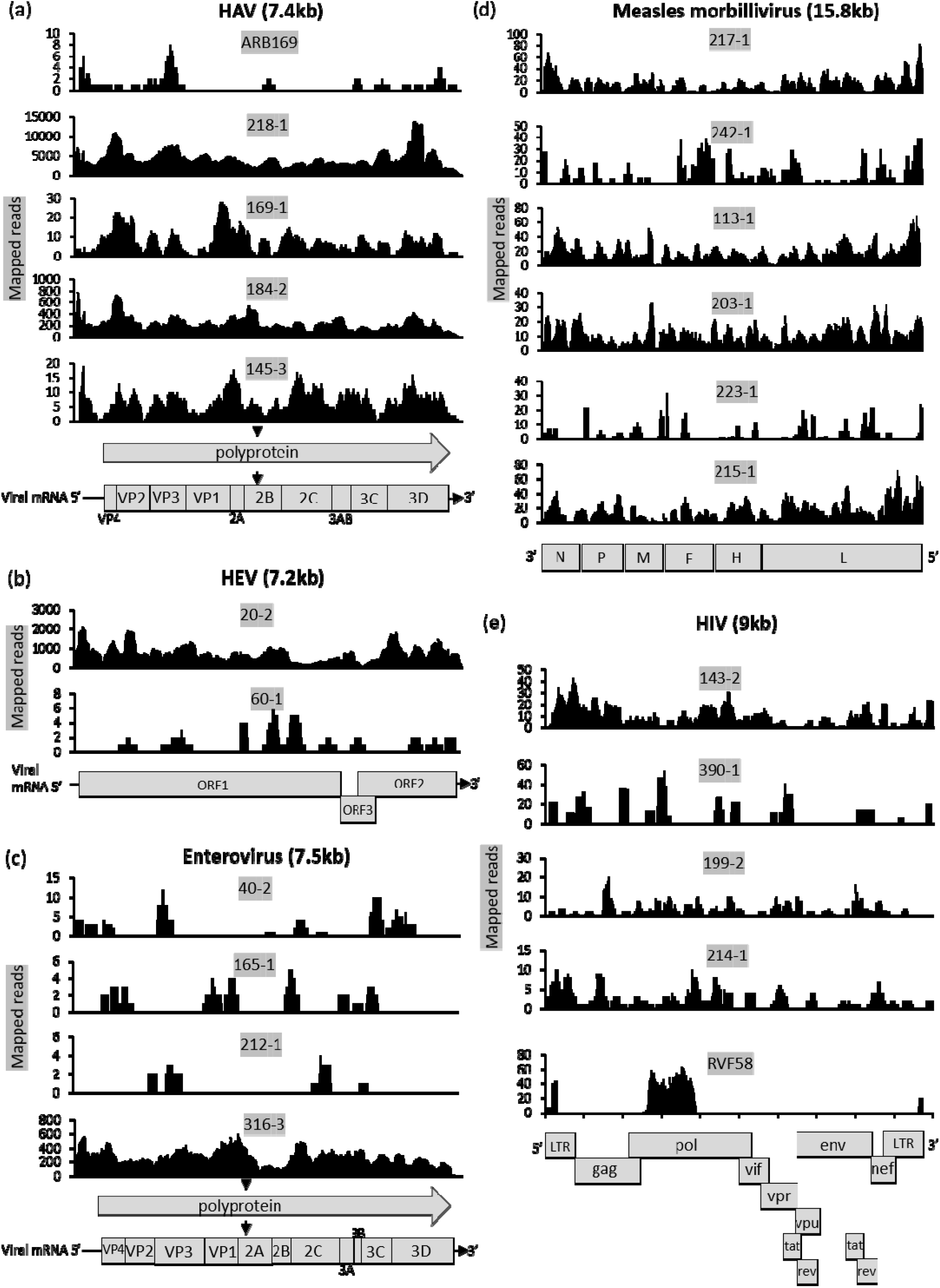
Other viral pathogens identified from patient samples. Coverage plots showing number of mapped reads (y-axis) against genome position (x-axis) for each patient, for (a) HAV, (b) HEV, (c) Enteroviruses including Echoviruses E3, E9/25, E4 and Rhinovirus C, (d) Measles virus; and (e) HIV-1. Reference genome drawings indicate depth for different regions along the sequence.

### Viruses associated with gastroenteritis

Gastrointestinal viruses were detected in plasma from 5 patients, including 4 enteroviruses (the echoviruses E3, E4 and E9 and Coxsackie B3 virus from patients 212-1, 316-3, 165-1, 195-1 respectively) and a fragment of a highly divergent rotavirus H-like virus in one patient (96-1) (**Figure 4c, S3f**).

### Viruses associated with respiratory infection

Respiratory viruses were detected in plasma from 9 patients and included six cases of measles morbillivirus (patients 215-1, 217-1, 242-1, 223-1, 113-1, 203-1), two cases of human parainfluenza virus 3 (patients 88-1, 114-1) and one case of rhinovirus C (patient 40-2) (**Figure 4c,d and S3b,c**). The measles virus sequences closely matched recently reported Ugandan B3 genomes. The rhinovirus C clustered with subtypes RV-C12 and RV-C44.

### Bloodborne viruses

The bloodborne viruses HBV (patient 192-1) and HIV-1 (patients RVF58, ARB176, 143-2, 390-1, 199-2, 214-1) may have occurred as acute or chronic infections, detected in a population with a high seroprevalence of both viruses. HBV was detected in a teenage patient (192-1) presenting with headache, anorexia and abdominal pain. It was most closely related to a sequence from Rwanda. HIV was identified as a co-infection in two patients, one with RVFV (RVF58) and another with YFV (ARB176) (**Figure 4e, S3a**); suggesting chronic infection. Patient 214-1 was infected with HIV-1 subtype A1, clustering with another Ugandan sequence. All other HIV-1 sequences were subtype D.

### Herpesviruses

Three patient samples (082-2, 185-2 and 143-2) contained human herpesvirus-6B (HHV6B) genome, the cause of roseola infantum, related to strains from USA, Uganda and DRC. Seven patients (187-2, 189-2, ARB244, ARB265, ARB454, ARB688 and HEV2) had evidence of cytomegalovirus (HHV5) in the bloodstream; this may have been acute or chronic infection. No varicella zoster virus (VZV) infections were detected.

### Viruses detected that are not known to be associated with disease

MNGS also detected viruses often found in human populations that have not been definitively associated with disease. These included the annelloviruses: Torque Teno virus (TTV), Torque Teno mini virus (TTMV), Torque Teno midi virus (TTMdV); and human pegiviruses (HPgV) (**Table S2**).

We also detected a second and novel rhabdovirus sequence in several AFI samples by *de novo* assembly (Adumi virus). This virus was likely to be a contaminant of blood collection tubes, as discussed in Supplementary Information.

## DISCUSSION

In this prospective cohort study of AFI in Uganda, we used mNGS to identify viral causes of acute undiagnosed fever in 210/1281 (16.4%) patients who tested negative after extensive screening for well-characterised pathogens, and an additional 23 samples obtained from outbreaks during the same time period. The cohort was prospectively recruited from patients presenting to three HCs in rural and urban Uganda. We detected viral pathogens in 35/233 (15.5%) patients, including 6/233 (2.6%) associated with VHF. Lack of clinical diagnosis of such viruses means that appropriate public health measures may not be taken to prevent onward transmission and appropriate treatment may not be given. The most commonly detected viruses reflected acute gastrointestinal or respiratory infection. The detection of active blood-borne virus infection reflects a chronic burden of disease and a need to widen treatment in the Ugandan population. Importantly, acute viral infection was invariably misdiagnosed as bacterial or malaria and treated with antibiotics or antimalarials. This carries an associated risk of antimicrobial drug resistance. We also noted vaccine-preventable infections in children (HAV, HBV, measles, Rotavirus) indicating the need for a more comprehensive vaccination schedule.

AFI is widespread in SSA and in the absence of specialised point-of-care diagnostics; clinical symptoms associated with different viruses may be indistinguishable. Even high-consequence VHFs such as Lassa fever and Ebola virus disease present with haemorrhage in less than 20% of cases ^19,20^. We identified CCHFV in a patient with undifferentiated fever who did not report contact with ticks or slaughtered animals, highlighting the limitations of current risk algorithms in identifying such patients in an endemic area. A high CCHFV seroprevalence has recently been reported in farming communities and domesticated animals in Uganda, suggesting that unrecognised infection may be common^21^.

We also detected the mosquito-borne infections RVFV, DENV and YFV; of which RVF can also be caused by contact with infected livestock. RVF is widespread across Africa and may cause a spectrum of illness from mild to severe. It is associated with haemorrhage and encephalitis in around 1% of human cases^22^. DENV is transmitted by *Aedes* mosquitoes and is endemic in Asia, South America and sub-Saharan Africa. It is relatively less well reported from the African continent, although studies suggest high seroprevalence^23,24^. Worldwide, there are an estimated 100 million clinical cases per year, of which around 5% progress to dengue shock syndrome or dengue haemorrhagic fever. We detected DENV-3 in this study, in keeping with previous reports of genotypes 1-3 on the African continent. YFV is endemic in Africa and South America and is transmitted by *Aedes* and *Haemagogus* mosquitoes. Severe disease with haemorrhage occurs in up to 12% of those infected^25^. An effective live vaccine exists for YFV and has recently been used to control outbreaks in Brazil and DRC. It is required for travellers to Uganda and other endemic regions, but the local unvaccinated population may remain vulnerable to infection.

We detected two rhabdoviruses in this study; LDV, a potential pathogen and Adumi virus, a likely contaminant. Rhabdoviruses are highly divergent and ubiquitous in host range, infecting plants, invertebrates and vertebrates. Human disease is well-described; rabies lyssavirus causes fatal neurological infection and Chandipura virus causes acute neurological disease. Other, less well described members have also been suggested as human pathogens, including Bas Congo virus that was associated with a single outbreak of haemorrhagic disease in the DRC. Additionally, studies have also identified rhabdoviruses in blood from healthy asymptomatic individuals^26,27^. LDV is an emerging pathogen and has only two historical documentations; in a child with hepatosplenomegaly and fever in Senegal in 1965 (sequenced in 2015) and in a middle-aged shipyard worker with fever and severe neurological symptoms in Wales in 1969, who had offloaded a ship inbound from Nigeria. LDV belongs to the genus *Ledantevirus* and closely related species have been found in rodents and bats. Neurological pathology has been described in rodents infected experimentally with the virus^28^. In our study, we demonstrated an immune response and the development of neutralising antibodies in the infected individual and a household contact who had similar symptoms at the time.

Vaccine-preventable diseases remain common in Uganda, even for viruses covered under routine immunisation, indicating insufficient coverage to generate herd immunity. Uganda has an active Expanded Programme of Immunization (EPI) against measles, HBV and rotavirus. YFV vaccination was introduced in October 2022. Vaccine coverage is lower in some neighbouring countries and this may increase the risk of imported infection. Measles vaccine in Uganda is often given as a single dose and booster vaccines during outbreaks. Such an approach may limit severe illness but has lower efficacy than the two-dose regimen used in other countries^29^.

We detected a viral pathogen in 15.5% of undiagnosed cases, however, the high detection of viruses associated with respiratory and gastrointestinal infection in plasma highlights the high burden that these diseases carry in Uganda. The inclusion of respiratory and stool samples (not available in this study) in future studies will likely reveal a much higher prevalence of infection. Additionally, the inclusion of serological assays would increase detection of infections outside the viraemic phase. We have likely under-estimated the burden of viral disease in this study for these reasons.

Viral metagenomics, although a powerful and sensitive method for unbiased genomic discovery is prone to limitations. Caution must be practiced when interpreting NGS data to exclude artefacts, which can arise from cross-contamination from the environment, within sequencing reagents and from other samples processed or sequenced together. In several studies, viral reads have subsequently been related to contamination within kits or from the environment. For example, Nextera XT reagents have been associated with detection of Merkel cell polyomavirus^30^. In this study, we found a novel rhabdovirus in several patient samples, most likely as a result of contaminated blood tubes with a virus of mosquito origin. This highlights the need for careful interpretation of mNGS data, confirmation by other methods and the development of standards and validation methods to interpret experimental information.

## CONCLUSIONS

Febrile illness is widespread in Uganda and a significant proportion of undiagnosed disease can be attributed to viral infections, several of which have pandemic potential and can cause high mortality and morbidity. Metagenomic next generation sequencing is a powerful tool to investigate the viral aetiology of AFI in high-risk populations and can be applied to develop low-cost clinical diagnostics. The finding of multiple highly pathogenic and poorly-described viruses in a single multi-site study in Uganda highlights the depth of the unknown with regard to circulating human viruses and the risk for onward transmission in a rapidly changing world.

## Data Availability

Anonymised data produced in the present study are available upon reasonable request to the authors. Clinically identifiable data will not be provided under restrictions for the use of the data provided by the UVRI REC.

## References

1. Carrington LB, Wills B. Lessons from history: viral surveillance in 1940s East Africa: Epidemiological notes on some viruses isolated in Uganda, GWA Dick, Transactions of the Royal Society of Tropical Medicine and Hygiene, 1953; 47 (1): 13–48. Trans R Soc Trop Med Hyg. 2018;112:413–4.

2. Mayanja MN, Mwiine FN, Lutwama JJ, Ssekagiri A, Egesa M, Thomson EC, et al. Mosquito-borne arboviruses in Uganda: History, transmission and burden. Journal of General Virology [Internet]. 2021 [cited 2023 Jan 17];102:001680. Available from: https://www.microbiologyresearch.org/content/journal/jgv/10.1099/jgv.0.001680

3. Mordecai EA, Ryan SJ, Caldwell JM, Shah MM, LaBeaud AD. Climate change could shift disease burden from malaria to arboviruses in Africa. Lancet Planet Health. 2020;4:e416–23.

4. Stoler J, Awandare GA. Febrile illness diagnostics and the malaria-industrial complex: a socioenvironmental perspective. BMC Infect Dis. 2016;16:1–9.

5. Murray CJL, Vos T, Lozano R, Naghavi M, Flaxman AD, Michaud C, et al. Disability-adjusted life years (DALYs) for 291 diseases and injuries in 21 regions, 1990–2010: a systematic analysis for the Global Burden of Disease Study 2010. The lancet. 2012;380:2197–223.

6. Prasad N, Murdoch DR, Reyburn H, Crump JA. Etiology of severe febrile illness in low-and middleincome countries: a systematic review. PLoS One. 2015;10:e0127962.

7. Crump JA, Morrissey AB, Nicholson WL, Massung RF, Stoddard RA, Galloway RL, et al. Etiology of severe non-malaria febrile illness in Northern Tanzania: a prospective cohort study. PLoS Negl Trop Dis. 2013;7:e2324.

8. Maze MJ, Bassat Q, Feasey NA, Mandomando I, Musicha P, Crump JA. The epidemiology of febrile illness in sub-Saharan Africa: implications for diagnosis and management. Clinical Microbiology and Infection. 2018;24:808–14.

9. Baba M, Logue CH, Oderinde B, Abdulmaleek H, Williams J, Lewis J, et al. Evidence of arbovirus coinfection in suspected febrile malaria and typhoid patients in Nigeria. The Journal of Infection in Developing Countries. 2013;7:51–9.

10. Jerome H, Taylor C, Sreenu VB, Klymenko T, Filipe ADS, Jackson C, et al. Metagenomic nextgeneration sequencing aids the diagnosis of viral infections in febrile returning travellers. Journal of Infection. 2019;79:383–8.

11. Buchfink B, Xie C, Huson DH. Fast and sensitive protein alignment using DIAMOND. Nat Methods. 2015;12:59–60.

12. Safonova Y, Bankevich A, Pevzner PA. dipSPAdes: assembler for highly polymorphic diploid genomes. Journal of Computational Biology. 2015;22:528–45.

13. Nguyen LT, Schmidt HA, von Haeseler A, Minh BQ. IQ-TREE: A Fast and Effective Stochastic Algorithm for Estimating Maximum-Likelihood Phylogenies. Mol Biol Evol [Internet]. 2015 [cited 2022 Aug 8];32:268–74. Available from: https://academic.oup.com/mbe/article/32/1/268/2925592

14. Kumar S, Tamura K, Nei M. MEGA: molecular evolutionary genetics analysis software for microcomputers. Bioinformatics. 1994;10:189–91.

15. Aricescu AR, Lu W, Jones EY. A time-and cost-efficient system for high-level protein production in mammalian cells. Acta Crystallogr D Biol Crystallogr. 2006;62:1243–50.

16. Hofmann K, Stoffel W. TMpred, Prediction of Transmembrane Regions and Orientation. 1993.

17. Whitt MA. Generation of VSV pseudotypes using recombinant ΔG-VSV for studies on virus entry, identification of entry inhibitors, and immune responses to vaccines. J Virol Methods. 2010;169:365–74.

18. Walker PJ, Firth C, Widen SG, Blasdell KR, Guzman H, Wood TG, et al. Evolution of genome size and complexity in the Rhabdoviridae. PLoS Pathog. 2015;11:e1004664.

19. Schieffelin JS, Shaffer JG, Goba A, Gbakie M, Gire SK, Colubri A, et al. Clinical illness and outcomes in patients with Ebola in Sierra Leone. New England journal of medicine. 2014;371:2092–100.

20. (CDC C for DC and P. Imported Lassa fever--New Jersey, 2004. MMWR Morb Mortal Wkly Rep. 2004;53:894–7.

21. Atim SA, Ashraf S, Belij-Rammerstorfer S, Ademun AR, Vudriko P, Nakayiki T, et al. Risk factors for Crimean-Congo Haemorrhagic Fever (CCHF) virus exposure in farming communities in Uganda. Journal of Infection. 2022;85:693–701.

22. Paweska JT. Rift Valley fever. In: Emerging Infectious Diseases. Elsevier; 2014. p. 73–93.

23. Simo FBN, Bigna JJ, Kenmoe S, Ndangang MS, Temfack E, Moundipa PF, et al. Dengue virus infection in people residing in Africa: a systematic review and meta-analysis of prevalence studies. Sci Rep. 2019;9:1–9.

24. Dieng I, Ndione MHD, Fall C, Diagne MM, Diop M, Gaye A, et al. Multifoci and multiserotypes circulation of dengue virus in Senegal between 2017 and 2018. BMC Infect Dis. 2021;21:1–11.

25. Johansson MA, Vasconcelos PFC, Staples JE. The whole iceberg: estimating the incidence of yellow fever virus infection from the number of severe cases. Trans R Soc Trop Med Hyg [Internet]. 2014 [cited 2023 Jan 23];108:482–7. Available from: https://academic.oup.com/trstmh/article/108/8/482/2765182

26. Stremlau MH, Andersen KG, Folarin OA, Grove JN, Odia I, Ehiane PE, et al. Discovery of novel rhabdoviruses in the blood of healthy individuals from West Africa. PLoS Negl Trop Dis. 2015;9:e0003631.

27. Grard G, Fair J, Chiu C, Leroy E. Bas-Congo virus: A novel rhabdovirus associated with acute hemorrhagic fever. In: Emerging infectious diseases. Elsevier; 2014. p. 13–24.

28. Woodruff AW, Ansdell VE, Bowen ET. Le Dantec virus infection in a patient who had not been to West Africa. Br Med J. 1977;2:1632.

29. Wichmann O, Hellenbrand W, Sagebiel D, Santibanez S, Ahlemeyer G, Vogt G, et al. Large measles outbreak at a German public school, 2006. Pediatric Infectious Disease Journal [Internet]. 2007 [cited 2023 Jan 17];26:782–6. Available from: https://journals.lww.com/pidj/Fulltext/2007/09000/Large_Measles_Outbreak_at_a_German_Public_School,.3.aspx

30. Asplund M, Kjartansdóttir KR, Mollerup S, Vinner L, Fridholm H, Herrera JAR, et al. Contaminating viral sequences in high-throughput sequencing viromics: a linkage study of 700 sequencing libraries. Clinical Microbiology and Infection. 2019;25:1277–85.

